# Misinterpretation of viral load in COVID-19

**DOI:** 10.1101/2020.10.06.20208009

**Authors:** Renan Lyra Miranda, Alexandro Guterres, Carlos Henrique de Azeredo Lima, Paulo Niemeyer Filho, Mônica R. Gadelha

## Abstract

Knowledge of viral load is essential for formulating strategies for antiviral treatment, vaccination, and epidemiological control of COVID-19. Moreover, patients identification with high viral load could also be useful to understand risk factors such as age, comorbidities, severity of symptoms and hypoxia to decide the need for hospitalization. Several studies are evaluating the importance of analyzing viral load in different types of samples, clinical outcomes and viral transmission pathways. However, in a great number of emerging studies cycle threshold (Ct) values by itself is often used as a viral load indicator, which may be a mistake. In this study, we compared tracheal aspirate with nasopharyngeal samples obtained from critically ill COVID-19 patients and demonstrate how the raw Ct could lead to misinterpretation of results. Further, we analyzed nasopharyngeal swabs positive samples and propose a method to reduce evaluation error that could occur from using raw Ct. Based on these findings, we show the impact that normalization of Ct values has on interpretation of viral load data from different biological samples from patients with COVID-19, transmission and lastly in relations with clinical outcomes.

**Importance:** In a pandemic, prevention of disease transmission is key. Reliable data for profiles of viral load are needed and important to guide antiviral treatment, infection control and vaccination. The differential expression of SARS-CoV-2 viral RNA among patient groups is a current topic of interest and viral load has been associated with a diversity of outcomes. However, in a great number of emerging studies cycle threshold (Ct) values by itself is often used as a viral load indicator, which may be a mistake. In this study, we compared tracheal aspirate with nasopharyngeal samples obtained from critically ill COVID-19 patients and demonstrate how the raw Ct could lead to misinterpretation of results. Based on these findings, we show the impact that normalization of Ct values has on interpretation of viral load data from different biological samples from patients with COVID-19, transmission and lastly in relations with clinical outcomes.

## Introduction

Besides investigating risk factors for mortality in hospitalized patients with coronavirus disease 2019 (COVID-19), such as older age, obesity, comorbidities, C-reactive protein (CRP), inflammatory cytokines, the impact of SARS-CoV-2 viral load in clinical outcomes could be extremely important(1–3). Moreover, reliable data for profiles of viral load are needed to guide antiviral treatment, infection control, epidemiological measures and vaccination. Several types of biological samples have been analyzed for the presence of SARS-CoV-2 viral RNA, as nasal swab, throat swab, sputum, rectal swab, vaginal swab, blood, placenta, human breastmilk, urine, among others(4, 5). Although in most of these types of samples the SARS-CoV-2 RNA was detectable, it’s not clear yet what is the pattern of viral load in these samples.

The differential expression of SARS-CoV-2 viral RNA among patient groups is a current topic of interest and viral load has been associated with a diversity of outcomes(5–8). The gold standard method to detect SARS-CoV-2 infection is the reverse-transcription quantitative PCR (RT-qPCR), which is based on the amplification of regions of viral RNA that have been reverse transcribed on each cycle of the reaction(9). The earlier the cycle that the fluorescence is detectable above a threshold, cycle threshold (Ct), indicates that the samples have a higher concentration of the target gene. In a great number of emerging studies Ct values by itself is often used as a viral load indicator. For example, raw Ct values were used to correlate viral load with a higher risk of intubation(6), to compare viral load between samples of nasopharyngeal (NPS) and oropharyngeal swabs (OPS)(10) and to investigate the relationship between Ct values and age range(8). Such application is common for the evaluation of viral loads of different type of viruses but high variability have been reported, often due to different equipment, PCR reagents, chemistry and standards used(11).

However, the amount of biological material retrieved by a swab could vary depending on the quality of the collection, thus a normalization attempt could prove useful when interpreting results(12). In the present work, we compared tracheal aspirate (TA) with nasopharyngeal samples (NPS) obtained from critically ill COVID-19 patients. Comparison on the relation between raw Ct value and ΔCt was used to demonstrate how the raw Ct could lead to misinterpretation of results. Further, we analyzed nasopharyngeal swabs positive samples and propose a method to reduce error that could occur from using raw Ct. Based on these findings, we explored the impact that Ct values normalization has on interpretation of obtained results of RT-qPCR data from biological samples of COVID-19 patients.

## Methods

### Samples

In this study, RT-qPCR data were obtained from 138 patients that tested positive for SARS-CoV-2. In total, there were 138 NPS samples, one from each patient, and 21 TA samples from intubated patients that were admitted in the intensive care unit, at Instituto Estadual do Cérebro Paulo Niemeyer, Rio de Janeiro, Brazil. TA samples were collected at the same day as NPS samples from each patient. The studies involving human participants were reviewed and approved by the ethical committee of Instituto Estadual do Cérebro Paulo Niemeyer (file number 3.997.619).

### RT-qPCR

The TaqMan™ RT-qPCR assays were performed in the QuantStudio 7™ Flex Real-Time PCR System (Applied Biosystems, Foster, CA, USA), directed to the nucleocapsid N gene regions (N1 and N2) of SARS-CoV-2 viral RNA (CDC assays for SARS-CoV-2 detection, manufactured by Integrated DNA Technologies – IDT, Iowa, USA). Thermal cycling was performed at 45 °C for 15 min for reverse transcription, followed by 95 °C for 2 min and then 45 cycles of 95 °C for 3 s and 55 °C for 30 s. A cycle threshold value less than 40 is interpreted as positive for SARS-CoV-2 RNA. In this assay, a RNase P gene region is used as an endogenous internal control for the analysis of biological samples. It is normally used to ensure the quality of the test, excluding the possibility of false negative due to the presence of eventual inhibitors or the quality and integrity of RNA samples(12). However, all human cells have a single-copy of the RNase P gene that encodes the mRNA moiety for the RNAse P enzyme. Therefore, their Ct values are associated with a range of input cell numbers in the RNA extraction(13). Thus, in order to evaluate possible variability in the amount of material retrieved from NPS and other specimen types we utilized RNase P as reference gene to normalize the input data.

### RT-qPCR normalization

When performing relative gene expression analysis of qPCR data, the first step known as Delta Ct (ΔCt) obtained by subtracting the reference gene Ct from target-gene Ct to account for input amount fluctuation that may occur(14). For this statement to be true, one needs to assume that amplification efficiency (E) would be ideally 100%. So we evaluated the E for both assays using standard curve analysis, since even though reported E is close to 100% it is of utmost importance to validate it with our laboratory setup(15). Then, we got ΔCt from our samples using RNaseP as a reference gene (ΔCt = Ct_N1_-Ct_RNaseP_). When comparing different sample types, TA and NPS, we used Ct and ΔCt on paired samples to check whether there was a difference in viral RNA load or in the amount of biological material. When evaluating RT-qPCR data of swabs we compared Ct and ΔCt and propose a method to reduce error that could occur from using raw Ct. We applied a formula that corrects the Ct values to achieve the closest relation to ΔCt values. This is a simple correction based on the formula proposed by Duchamp et al., 2010(16). They used this formula to correct influenza A viral load per sample, calculating a Ct value modified according to the ratio of sample RNase P and mean RNase P Ct values ([sample influenza A Ct value x sample RNaseP Ct value/mean RNaseP Ct value]).

### Statistical analysis

All data analysis was performed with the GraphPad Prism 6 (GraphPad Software Inc., USA). Data were expressed as mean ± standard deviation. The Student t-test was used for comparison between two groups. Spearman correlation was used to compare the relationship between N1 Ct and ΔCt. Differences were considered to be significant at a level of P < 0.05.

## Results

### Uncorrected Ct values and misinterpretation of viral load

Before analyzing results, we evaluated E of the TaqMan™ assay from CDC kit: E of 100.177% for the N1 assay (R^2^ = 0.999, slope = −3318, error = 0.03); 98.322% for the N2 assay (R^2^ = 0.997, slope = −3363, error = 0.045); and 107.274% (R^2^ = 0.997, slope = −3159, error = 0.045) for the RNase P assay. Then, we performed the following tests using only N1 as a viral target since it had a better E. When comparing 21 paired samples of TA and NPS: TA samples have a lower N1 Ct value than NPS samples (P < 0.001), meanwhile having lower RNase P Ct values as well (P < 0.05) (Figure 1A); however, if we compare the ΔCt values from the paired samples we get that there is no difference between TA and NP samples (P = 0.859) (Figure 1B). It is important to note that for one patient the NP sample was negative for SARS-CoV-2 and the TA sample was positive (N1 Ct = 34). The difference in Ct values having similar ΔCt values indicates that the higher concentration of viral RNA in TA samples is a consequence of a higher concentration of total RNA.

**Figure 1.**
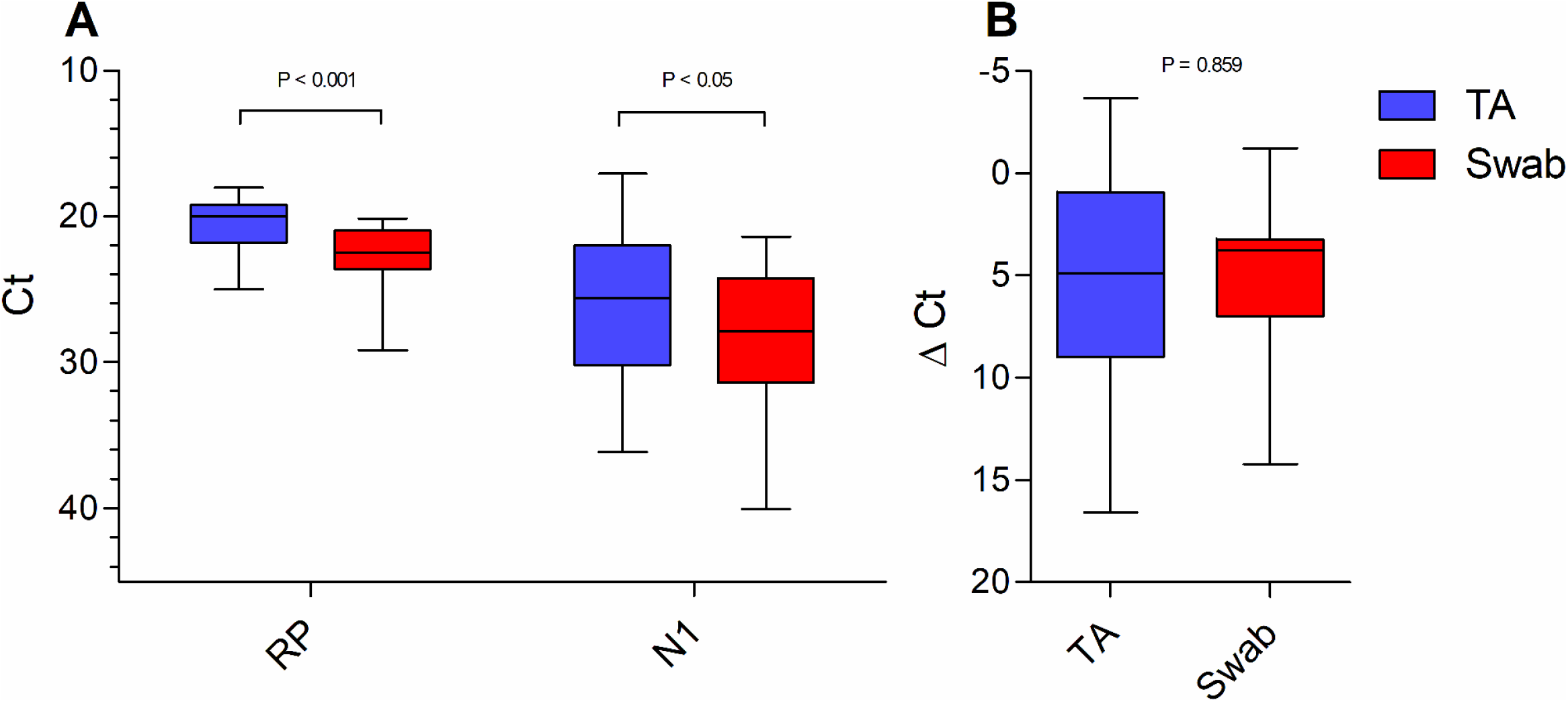
Comparison between nasopharyngeal swabs and tracheal aspirates for SARS-CoV-2 detection. **(A)** N1 and RP Ct values of NPS X TA samples (**N1**: P < 0.05, TA = 25.6 (17.04 – 36.11) / 26.13 ± 4.99 and NPS = 27.87 (21.37 – 31.36) / 28.22 ± 4.54; **RP**: P*< 0.001, TA = 19.94 (18.02 – 24.98) / 20.49±1.76 and NPS = 22.47 (20.11-29.16) / 22.61 ± 2.09). **(B)** ΔCt (N1 – RP) of NPS X TA samples (P = 0.859, TA = 4.90 (−3.71 – 16.59) / 5.64 ± 5.65 and NPS = 3.74 (−1.21 – 14.21) / 5.06 ± 3.91). Data are expressed as media (min – max)/ mean ± standard deviation, statistical difference was evaluated by paired T test. (RP = RNAse P, NPS = nasopharyngeal samples, TA = tracheal aspirate).

### Discrepancy between uncorrected Ct values and ΔCt values

In order to demonstrate the discrepancy that can arise when comparing results of N1 Ct and ΔCt we plotted those values obtained from 138 NPS positive samples. The summary of statistics is as follows: N1, mean = 25.31/ StdD = 5.47; RP, mean = 24.99/ StdD = 2.10; ΔCt, mean = 0.32/ StdD = 5.31. Even though we do have a correlation between those values (R = 0.94) it could provide a misleading result. On the X axis a variation of 1 ΔCt from −3 to −2 includes 11 samples that have N1 Ct values ranging from 18.61 to 25.5. Interestingly, if we look at the ΔCt of these min and max N1 Ct values within this range we get −2.1 and −2.21, respectively (Figure 2A). If uncorrected Ct values were to be used as a measure of viral load difference between those samples we would get a difference of 6.89 cycles, which would correspond roughly for a difference of 118 times more viral RNA present in the sample with lower Ct, meanwhile if we apply the fold change formula (2^-ΔΔCt^) to compare the same samples we would get a fold change of 1.08.

**Figure 2.**
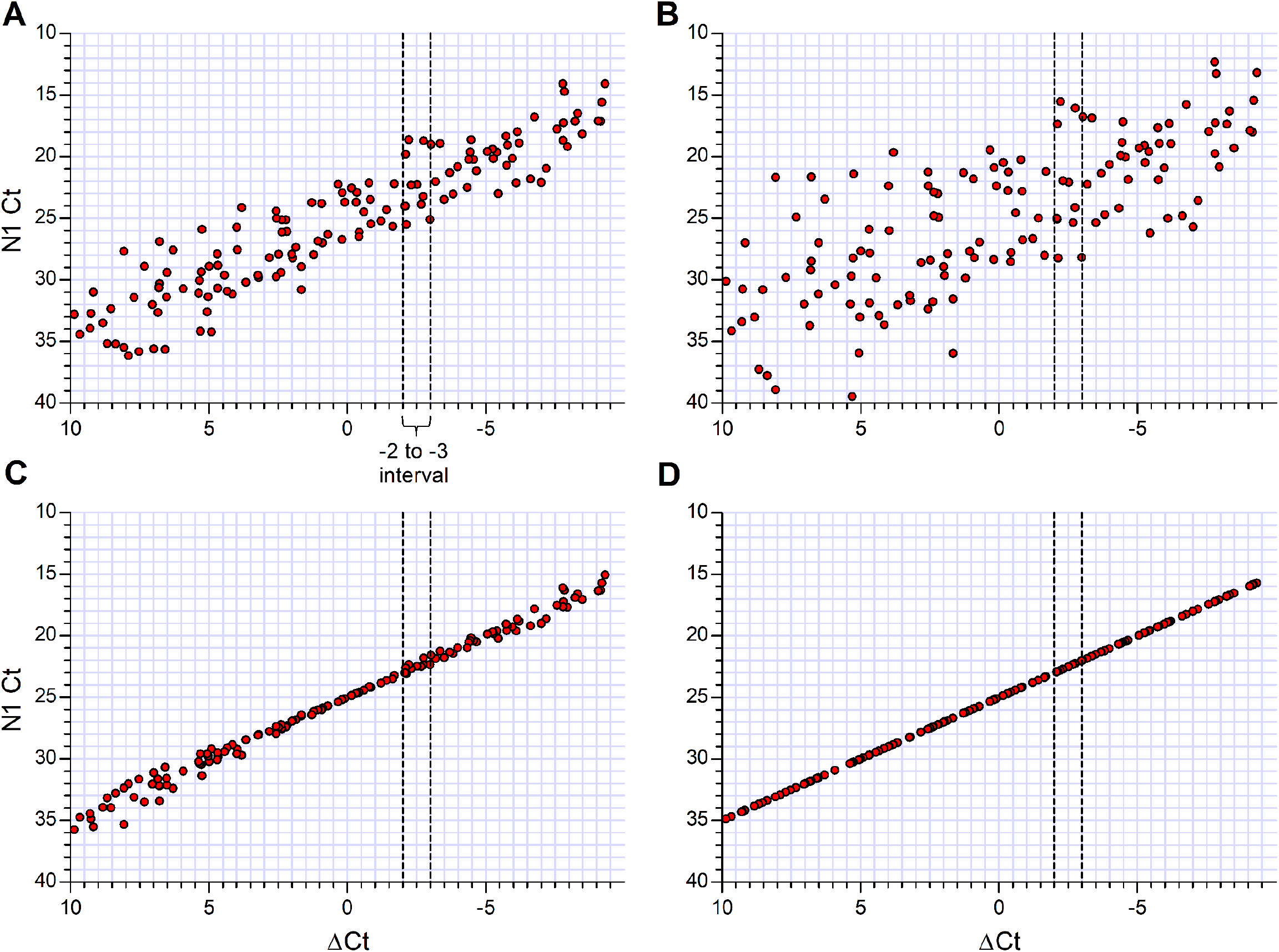
N1 Ct X ΔCt using different corrections. **(A)** No correction. **(B)** Correction proposed by Duchamp et al., 2010: Ct = Ct_N1_* Sample Ct_RNaseP_ /mean Ct_RNaseP._ **(C)** Modification on method proposed in B: Ct = Ct_N1_* mean Ct_RNaseP_/Sample Ct_RNaseP._ **(D)** Method with direct relation to ΔCt variation Ct = Ct_N1_ – (Sample Ct_RNaseP_ – mean Ct_RNaseP_).

### Reducing the discrepancy between Ct and ΔCt

We then applied a formula to correct Ct values based on the RNase P mean Ct (Ct_N1_* sample Ct_RNaseP_ /mean Ct_RNaseP_), as proposed by Duchamp et al., 2010(16), however as can be observed on Fig. 2B this method further increase the distance in Ct values of samples that had similar ΔCt values (a difference of cut-off cycle threshold values of 12.70), which is an undesirable effect. We also observed a decrease in the correlation between those values (R=0.76). We modified this formula trying to decrease the discrepancy of original Ct values of samples with similar ΔCt values, since the discrepancy Ct values in similar ΔCt values are result of differences in the amount of biological material used in the input. A modification of the formula was used (Ct_N1_* mean Ct_RNaseP_/sample Ct_RNaseP_). After the adjustment the Ct value of the min goes from 18.61 to 22.9 and max from 25.5 to 23.7, now they have a difference of 0.8 cycles that would be somewhere around 1.74 times more viral RNA. This adjusted Ct has even stronger correlation to the original ΔCt value achieving a spearman rank of 0.99 (Figure 2C).

Even reducing the discrepancy and increasing the correlation, we observed that Ct values <20 and >30 were not adjusted in a similar way to intermediate values. We apply a third formula, where we get the difference between sample RNase P Ct and mean RNase P Ct, and then subtract it from sample N1 Ct (Ct_N1_ – (Sample Ct_RNaseP_ - mean Ct_RNaseP_)). With this, all Ct values become directly related with ΔCt values, yielding a correlation value of R=1 (Figure 2D).

## Discussion

The impact of the pandemic on our society has increased the demand for quick responses and solutions, pushing the adaptation of sample collection due to shortage of materials like the use of nasopharyngeal swabs or oropharyngeal swabs(17). However, the importance of systematic validation remains, although the potentially misleading effects of using raw data, inappropriate references for normalization or even non-standardization are being widely considered. Consequently, real-time RT-qPCR data obtained in diagnostic of COVID-19 are being used in many molecular analyzes especially for viral load determination. Due to the diversity of sample types, variations in the quantities of imputing material, commercial detection kits and experimental conditions, it becomes impossible to control all parameters involved in COVID-19 diagnosis. Therefore, reference data, normalization, quantification process efficiency must be considered when we use data from real-time RT-qPCR analysis.

In our study, we demonstrated that considering the Ct values without any correction, TA samples have significantly (P < 0.001) more SARS-CoV-2 viral RNA than the NP samples. However, we can clearly see that RNAse P Ct values are significantly different (P<0.05), indicating that tracheal aspirates have higher amounts of biological material when compared to swabs. In short, when we perform the extraction of total RNA from, for example, 200 μL tracheal aspirate, it does not correspond to 200 μL swab. Even though this method would not provide actual viral RNA quantification it would be enough to show how the use of raw Ct can be misleading, and it is easy to apply even on a diagnostic setup. The study by Liu and colleagues(7) is one of the few that uses ΔCt. They observed that the ΔCt values of severe cases were significantly lower than those of mild cases at the time of admission. They indicated that mean viral load of severe cases was around 60 times higher than that of mild cases, suggesting that higher viral loads might be associated with severe clinical outcomes. However, one of the most cited studies on viral load (>860 citations) used only raw data of Ct values (18).

Pujadas and collaborators(19) showed an independent relation between high viral load and mortality. These authors reinforced the importance of transforming qualitative testing into a quantitative measurement of viral load will assist clinicians in risk-stratifying patients and choosing among available therapies and trials. However, Wang and collaborators(10) evaluated nasopharyngeal (NPS) and oropharyngeal swabs (OPS) specimens collected from 120 patients with confirmed COVID-19. They found mean Ct value (uncorrected) for NPS of 37.8 that was significantly lower than that of OPS 39.4, indicating that the SARS-CoV-2 load was significantly higher in NPS specimens than OPS. If sample concentration were to be taken into account a different conclusion could have been drawn from such comparison. Thus, it is extremely important to have an internal control for a human reference gene when comparing samples.

Heald-Sargent et al.(5) describe that levels of viral nucleic acid in NPS are significantly greater in children younger than 5 years, when compared with older children. Authors report that young children younger than 5 years and older children aged 5 to 17 years, had median cut-off cycle threshold (Ct) values (uncorrected) of 6.5 and 11 respectively. We demonstrated that within a range as far as 7 cycles in Ct for a viral marker samples could actually have a difference of only 0.1 cycles when ΔCt is taken into consideration. A multicentric study has demonstrated that viral load estimations for several viruses can vary considerably between different laboratories since there is no standardized required resources(11). Fernandes-Monteiro and collaborators demonstrated that serum samples tested for yellow fever had small variation in RNase P, even though there was significant difference in viral load between samples(13). For other sample types, like NPS, RNase P Ct could vary depending on the quality of sample and efficiency of acquisition(12).

Previously, Wang and collaborators (4) investigated the biodistribution of RNA viral among different types of biological samples, including bronchoalveolar lavage fluid, fibrobronchoscope brush biopsy, sputum, feces, blood, urine, among others. They evaluated 1070 specimens collected from 205 patients with COVID-19 and observed that Ct values (uncorrected) of all specimen types were higher than 30, except for nasal swabs with a mean Ct values of 24.3 (range of 16.9 to 38.4). However, without a correction in the Ct values it is not possible to confirm these differences. Recently, Vivanti and collaborators(5) demonstrated the transplacental transmission of SARS-CoV-2 in a neonate born to a mother infected in the last trimester and presenting with neurological compromise. In this study the authors detected SARS-CoV-2 RNA in amniotic fluid, vaginal and rectal swab, blood and NPS and call attention for a very high viral load in placenta. However, an important point is that different types of biological samples have different concentrations in number of cells and particles. For example, the human placenta is composed by a complex of fetal cells and is characterized by a close association between fetal-derived trophoblasts and the maternal tissues that they come into contact(20). Moreover, the complex composition of some samples types include proteins, fats, humic acid, phytic acid, Immunoglobulin G, bile, calcium chloride, EDTA, heparin and ferric chloride, and many of them have been recognized as PCR inhibitors(21).

Real-time RT-PCR has become a common technique, it is in many cases the main method for measuring the presence of viral RNA due to its sensitivity and a high potential for accurate quantification. Despite RT-qPCR inability to differentiate between infective and non•infective (antibody-neutralized or dead) viruses, using an estimative of viral RNA load remains plausible for clinical hypotheses formulation. The evaluation of infectiveness of a sample is not a simple procedure since virus isolation in cell culture of SARS-CoV-2 should be conducted in a Biosafety Level 3 (BSL-3) laboratory(9). To achieve this, however, appropriate normalization strategies are required to control for experimental error introduced during the multistage process required to extract and process the viral RNA. We agree that the ideal approach is to use Standard Curve Method using an endogenous control. In this method, for quantification normalized to an endogenous control, standard curves are prepared for both the target and the endogenous reference. For each experimental sample, the amount of target and endogenous reference is determined from the appropriate standard curve. However, this method has a high cost since standard curves need to be in all experiments.

Lastly, we are proposing a formula that is able to perform a perfect correlation between the corrected Ct values and ΔCt values, allowing new studies to use these corrected Ct values to calculate the number of viral copies. In conclusion, we have demonstrated that, overall, TA samples have more total RNA than NPS, even though there was no difference in viral load. Thus, if a reference gene is taken into consideration when analyzing NPS, samples that initially would be considered to have different viral loads by raw Ct comparison would actually have the same viral load. Thus, when comparing samples the use of reference gene is extremely important before drawing conclusions related COVID-19 viral load.

## Data Availability

All information is contained in the manuscript.

## Acknowledgements

This work was supported by grants from Fundação de Amparo a Pesquisa do Estado do Rio de Janeiro (FAPERJ).

## Author contributions

In terms of contributions authors RLM, AG and CHAL, worked directly with samples, performed the literature search, prepared the figures, interpreted the date and wrote the manuscript draft; MG and PNF participated in this work design, discussion of results and manuscript preparation. All authors critically reviewed the manuscript for important intellectual content and approved it in its final version.

